# COVID-19 vaccine effectiveness among South Asians in Ontario: A test-negative design population-based case-control study

**DOI:** 10.1101/2023.12.08.23299660

**Authors:** Rahul Chanchlani, Baiju R. Shah, Shrikant I. Bangdiwala, Russ de Souza, Jin Luo, Shelly Bolotin, Dawn ME Bowdish, Dipika Desai, Scott A Lear, Mark Loeb, Zubin Punthakee, Diana Sherifali, Gita Wahi, Sonia S. Anand

## Abstract

**Objectives:** To: 1) evaluate the effectiveness of COVID-19 vaccines among South Asians living in Ontario, Canada compared to non-South Asians, and 2) compare the odds of symptomatic COVID-19 infection and related hospitalizations and deaths among non-vaccinated South Asians and non-South Asians.

**Design:** Test negative design study

**Setting:** Ontario, Canada between Dec 14, 2020 and Nov 15, 2021

**Participants:** All eligible individuals >18 years with symptoms of COVID-19 and subdivided by South Asian ethnicity versus other, and those who were vaccinated versus non-vaccinated.

**Main Outcome measures:** The primary outcome was vaccine effectiveness as defined by COVID-19 infections, hospitalizations, and deaths, and secondary outcome was the odds of COVID-19 infections, hospitalizations, and death comparing non-vaccinated South Asians to non-vaccinated non-South Asians.

**Results:** 883,155 individuals were included. Among South Asians, two doses of COVID-19 vaccine prevented 93.8% (95% CI 93.2, 94.4) of COVID-19 infections and 97.5% (95% CI 95.2, 98.6) of hospitalizations and deaths. Among non-South Asians, vaccines prevented 86.6% (CI 86.3, 86.9) of COVID-19 infections and 93.1% (CI 92.2, 93.8) of hospitalizations and deaths. Non-vaccinated South Asians had higher odds of symptomatic SARS-CoV-2 infection compared to non-vaccinated non-South Asians (OR 2.35, 95% CI 2.3, 2.4), regardless of their immigration status.

**Conclusions:** COVID-19 vaccines are effective in preventing infections, hospitalizations and deaths among South Asians living in Canada. The observation that non-vaccinated South Asians have higher odds of symptomatic COVID-19 infection warrants further investigation.

**What is already known?:** Some ethnic communities, such as South Asians, were disproportionately impacted during the COVID-19 pandemic. However, there are limited data on COVID-19 vaccine efficacy among this high-risk ethnic group.

**What this study adds?:**

- In this large population-based study including close to 900,000 individuals in Canada, we show COVID-19 vaccines are effective in preventing symptomatic SARS CoV-2 infections, hospitalizations and deaths among both South Asians and non-South Asians.
- We also demonstrate that, among non-vaccinated individuals, South Asians have higher odds of COVID-19 infection, and an increased risk of COVID-19 hospitalizations and deaths compared to non-South Asians.

## Background

South Asians, or people who originate from the Indian subcontinent, are the largest non-white and fastest growing ethnic group in Canada. Some ethnic communities, such as South Asians, were disproportionately impacted during the COVID-19 pandemic(1). A recent population-based cohort study of 17 million adults from the United Kingdom found that South Asians were at increased risk of testing positive for SARS-CoV-2 and of COVID-19-related hospitalizations, ICU admissions, and deaths (2).

In Canada, people living in ethnically diverse neighbourhoods were shown to have three times higher rates of COVID-19 infection, four times higher rates of hospitalization and intensive care unit admissions, as compared to neighbourhoods with little ethnic diversity in 2020 (3). In the Peel Region in Ontario, between April 2020 and January 2021, racialized people accounted for 63% of the population, yet comprised 81% of COVID-19 cases, with South Asians being among the most commonly affected.(4) Similarly, in a cross-sectional analysis, our group showed South Asians living in Southwestern Ontario had a relatively high seropositivity (23.6%) of COVID-19 infection during wave 3 of the pandemic(5).

Vaccines are effective in reducing the risk of COVID-19 infection and associated hospitalizations and deaths, as shown by Phase 3 randomized clinical trials. (6-9). However, there is significant disparity in vaccine uptake among ethnic groups, especially South Asians (10-12). Approximately 25% of people living in the Indian subcontinent are vaccine hesitant due to cultural or religious reasons, concomitant comorbidities, low health literacy, receipt of misinformation on social media, and due to the influence of their peers (13). Furthermore, in COVID-19 vaccine trials, “Asian” participants in which South Asians would be included, were substantially underrepresented (<5%) (9, 14), and therefore, there are sparse data on effectiveness of COVID-19 vaccines among South Asians. All of these factors may limit vaccine confidence and therefore uptake amongst the South Asian population in Canada.

To address these concerns, we conducted a series of studies (5, 15), including this population-based study using health administrative databases among South Asians living in Ontario, Canada. The specific objectives were to: 1) evaluate the effectiveness of COVID-19 vaccines among South Asians living in Ontario, Canada compared to non-South Asians, and 2) compare the odds of symptomatic COVID-19 infection and related hospitalizations and deaths among non-vaccinated South Asians and non-South Asians.

## Methods

This study is reported in accordance with the Reporting of studies Conducted using Observational Routinely collected health Data (RECORD) guidelines (16). The public was not involved with the design, conduct, reporting, or dissemination plans of this study.

### Study population, setting and design

Using the methods described by Chung et al (17), we conducted a population-based study with a test negative design in which we included all individuals 18 years and older living in Ontario who had symptoms consistent with COVID-19. Test-negative designs are often used for vaccine efficacy studies, and are a special case of a case-control study in which the controls are subjects undergoing the same tests for the same reasons, but who test negative instead of positive as cases (18). All Ontarians who were tested by PCR for SARS-CoV-2 between December 14, 2020 and Nov 15, 2021, were eligible. We excluded those without OHIP (Ontario Health Insurance Plan) coverage (public health insurance) and those living in long-term care homes. We restricted the analysis to individuals who had at least one relevant COVID-19 symptom at the time of testing (17).

Symptomatic individuals who tested positive at least once for SARS-CoV-2 were considered as cases. Individuals who were symptomatic but were negative on all tests for SARS-CoV-2 during the study period were considered as controls. For cases, the index date was the date of specimen collection for their positive test (or one selected at random, if they had multiple positive tests), and for controls, the date of a randomly selected negative test result was considered as the index date.

Further, this cohort was divided into four sub-samples based on ethnicity and vaccination status, as follows: South Asian vaccinated, South Asian non-vaccinated, non-South Asian vaccinated, and non-South Asian non-vaccinated. Only those who had received both vaccine doses before the PCR testing and were 7 or more days after the second dose were considered as vaccinated.

#### Data sources

We obtained information regarding COVID-19 vaccination status, including vaccine product, date of administration, and dose number, from COVaxON, a centralized COVID-19 vaccine information system in Ontario. Data on laboratory-confirmed SARS-CoV-2 infection was detected by real-time reverse transcription polymerase chain reaction (RT-PCR) collected from the Ontario Laboratories Information System (OLIS) for both individuals who tested positive and individuals who tested negative. We obtained information on the clinical course of cases from the Public Health Case and Contact Management system (CCM). Ethnicity was determined using the ETHNIC database, which uses a validated last name algorithm to classify individuals into South Asians and non-South Asian groups (19). By design, this algorithm has high specificity for South Asian names, but low sensitivity. Immigration, Refugee and Citizenship Canada’s Permanent Resident Database was used to determine the immigration status and duration. Demographic data were obtained from the Registered Persons Database (RPDB). Hospitalization data were obtained from the Discharge Abstract Database [using International Classification of Diseases, Ninth and Tenth Revision (ICD-9 and ICD-10) codes], and diagnostic and fee codes from physician billing claims were obtained from the OHIP database. These datasets were linked using unique encoded identifiers and analyzed at ICES (*formerly known as* Institute for Clinical Evaluative Sciences).

#### Outcomes ascertainment

The primary outcome was symptomatic SARS CoV-2 infection and COVID-19 associated hospitalizations and deaths among vaccinated South Asians and non-South Asians. COVID-19-related hospitalization was defined as a positive test result which occurred within 14 days before or three days after admission. COVID-19-related death was identified as a positive test result which occurred within 30 days before death or within seven days postmortem (17).

**Secondary outcomes** included the odds of symptomatic SARS CoV-2 infection among non-vaccinated South Asians and non-South Asians; and the risk of COVID-19 associated hospitalization or death in COVID-19 infected non-vaccinated South Asians and non-vaccinated non-South Asians.

#### Covariates

Demographic variables included age, sex, neighborhood income quintile (by postal code), rural status (community <10,000 persons), immigration status, time since immigration and reason for immigration (economic, refugee and other). Pre-existing co-morbid conditions on ICES derived cohorts such as hypertension, diabetes, history of asthma, chronic obstructive pulmonary disease (COPD), cancer, CKD (chronic kidney disease), immunocompromised status, dementia/frailty, CHF (congestive heart failure), TIA/Stroke, cardiac ischemia, arrythmia were ascertained using various diagnostic and procedural codes (Supplementary Table 1).

### Statistical analysis

Descriptive statistics and standardized differences were used to compare the 4 groups. We used multivariable logistic regression models to calculate the odds ratio (and 95% confidence interval) for outcomes after adjusting for covariates (age, sex, rural/urban, neighborhood income quintile and any comorbidity). We then calculated the vaccine effectiveness for both symptomatic SARS CoV2 infection and for COVID-19 related hospitalizations and deaths, using the following formula: (1 – odds ratio of the outcome among vaccinated versus non-vaccinated individuals x 100%), in both the South Asian and non-South Asian populations. We also compared the odds of COVID-19 infection and the risk of COVID-19 related hospitalization and deaths among South Asian non vaccinated vs. non-South Asian non-vaccinated individuals. Stratified analyses were conducted using immigration status (non-immigrants, recent immigrants [<10 years] and non-recent immigrants [>/=10 years]), and, among immigrants only, the reasons for immigration (economic, refugee, and family/others). All analyses were conducted using SAS version 9.4 (SAS Institute, Cary, NC). Tests were two sided, with P<0.05 considered as significant.

## Results

A total of 883,155 individuals who had PCR test for SARS-CoV-2 between 14 December 2020 and November 15, 2021, were identified. Among them, 126,016 were cases and 757,139 were controls. (Supplemental Figure 1) Supplementary Table 2 depicts the cohort creation process. Baseline characteristics and comorbidities of the cases and controls are shown in Supplemental Tables 3 and 4.

The overall cohort was further divided into 4 subgroups as shown in Table 1. South Asians included in this analysis were younger, more frequently male, and less likely to live in rural communities compared to the non-South Asian group. More than half (54.8%) of South Asians resided in LHIN (local health integrated networks) 5 and 6 regions (i.e., Central West including city of Brampton, and Mississauga Halton). Notably, South Asians had an overall lower prevalence of comorbid conditions compared to non-South Asians, except for diabetes mellitus (Table 2).

**Table 1:**
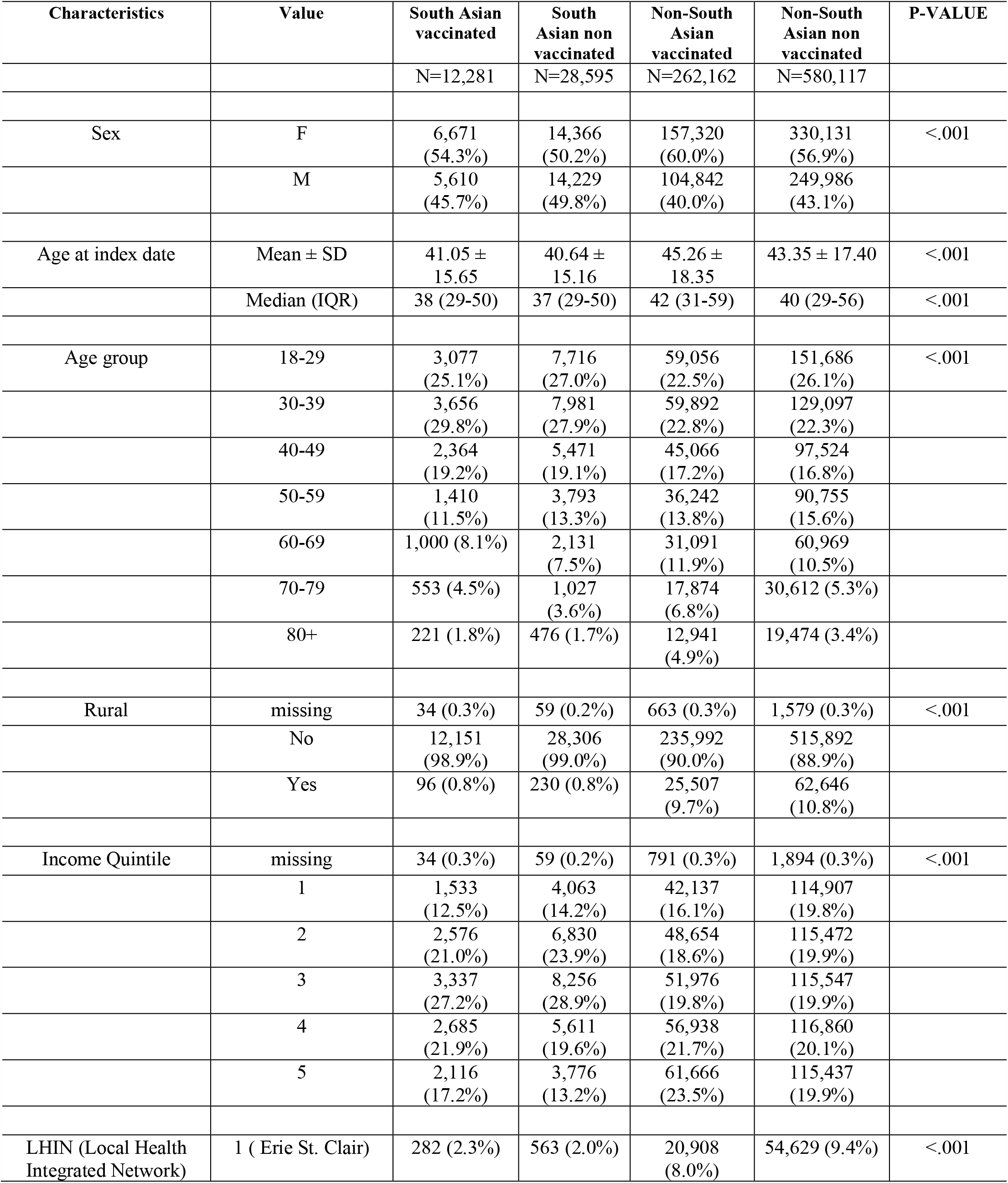
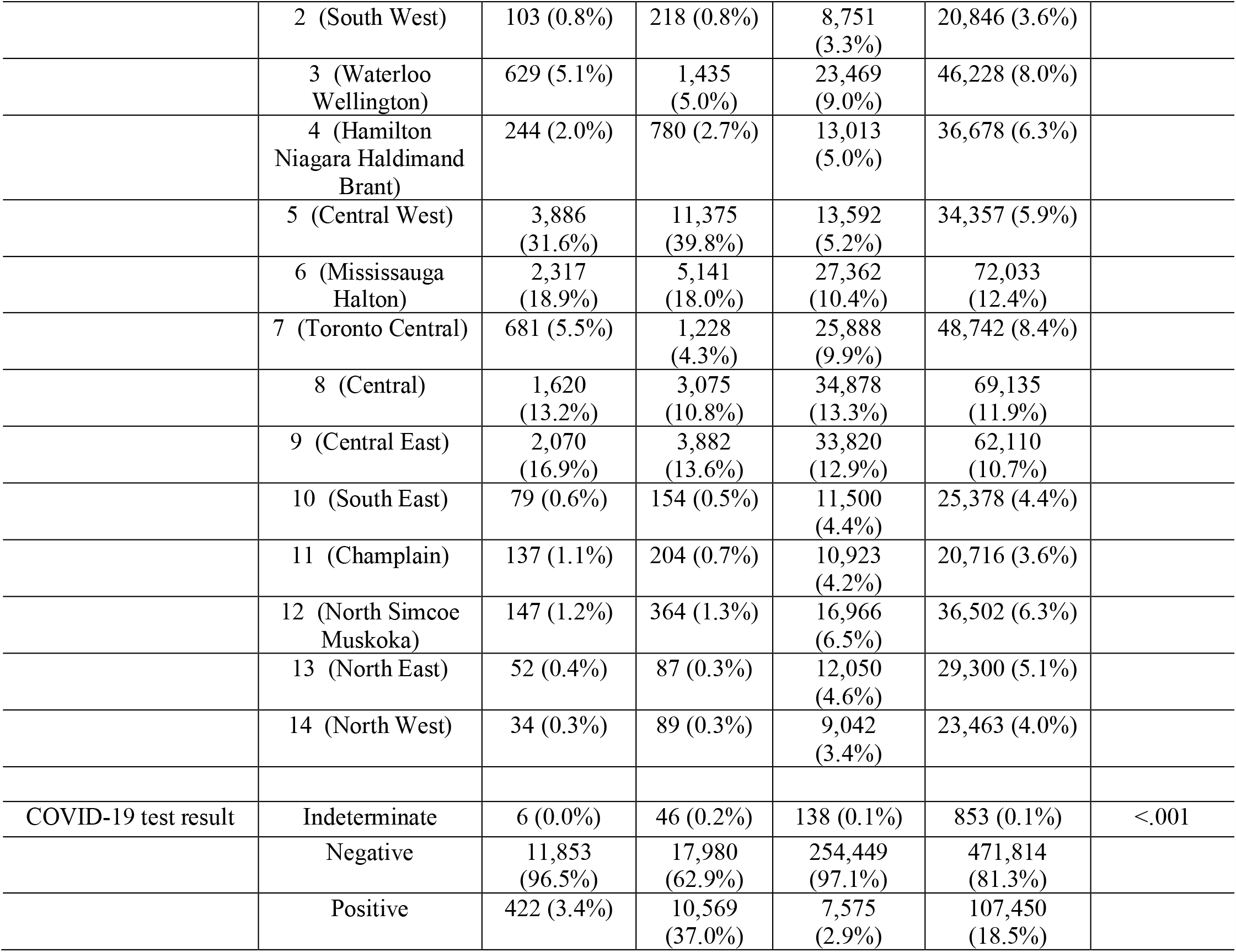
Baseline characteristics of the overall cohort stratified by ethnicity and vaccination status.

**Table 2:**
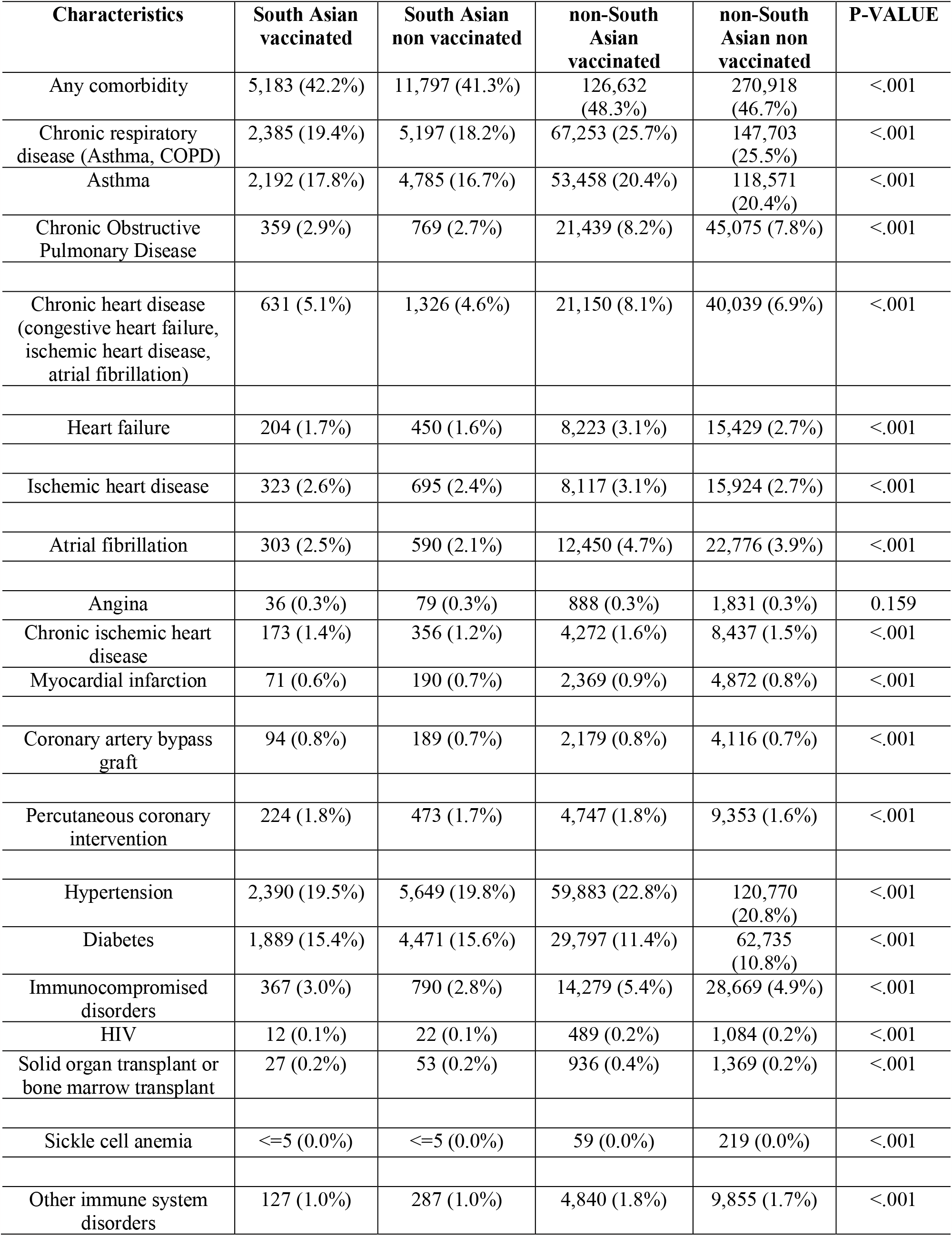
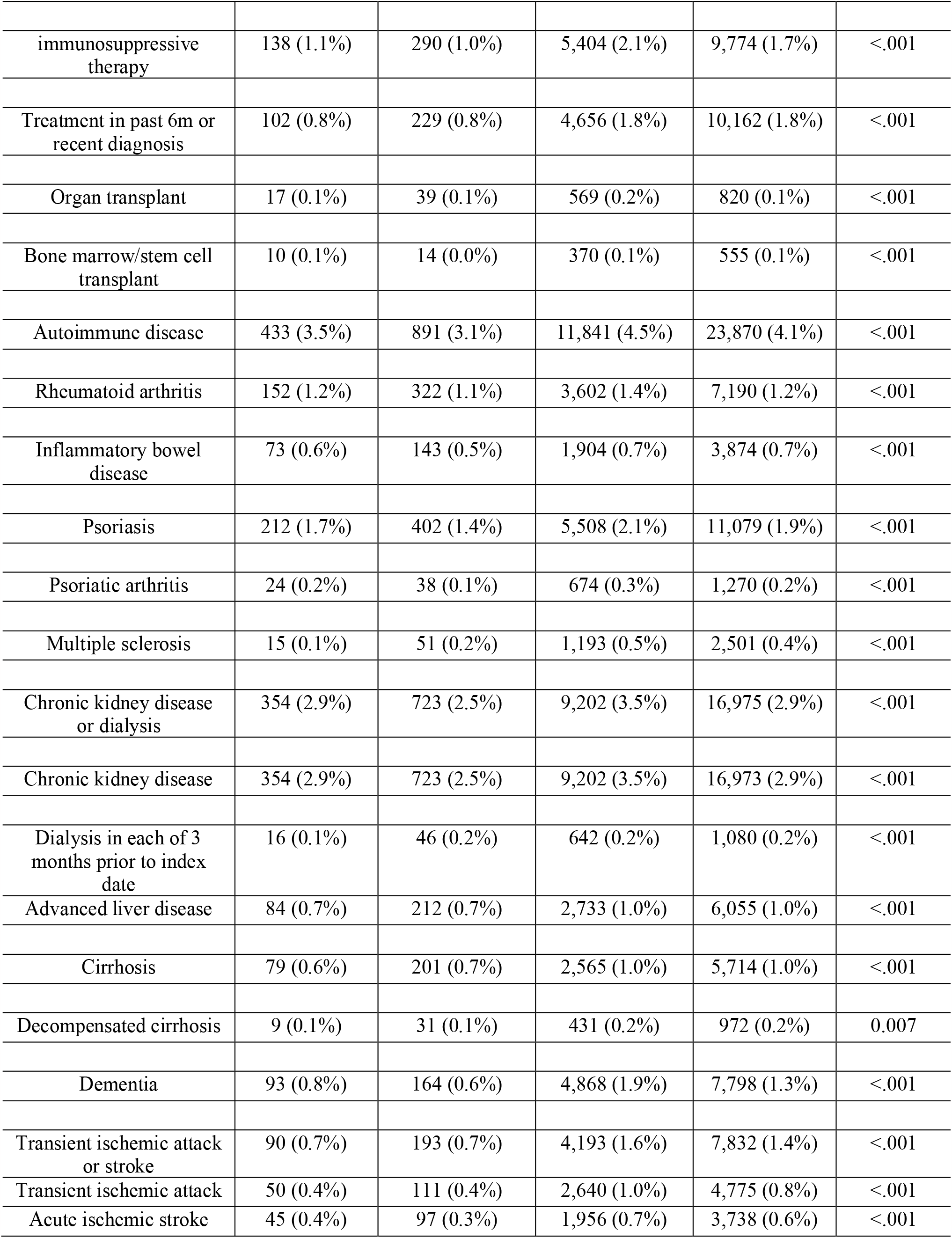
Baseline comorbidities of the overall cohort stratified by ethnicity and vaccination status.

### Vaccine effectiveness

Among South Asians, 2 doses of COVID-19 vaccine were 93.8% (95% CI 93.2, 94.4) and 97.5% (95% CI 95.2, 98.6) effective in preventing SARS Co-V-2 infection and related hospitalizations or deaths, respectively. Among non-South Asians, vaccine effectiveness was lower at 86.6% (CI 86.3, 86.9) and 93.1% (CI 92.2, 93.8) for SARS Co-V-2 infection and related hospitalizations or deaths, respectively. (Table 3)

**Table 3:**
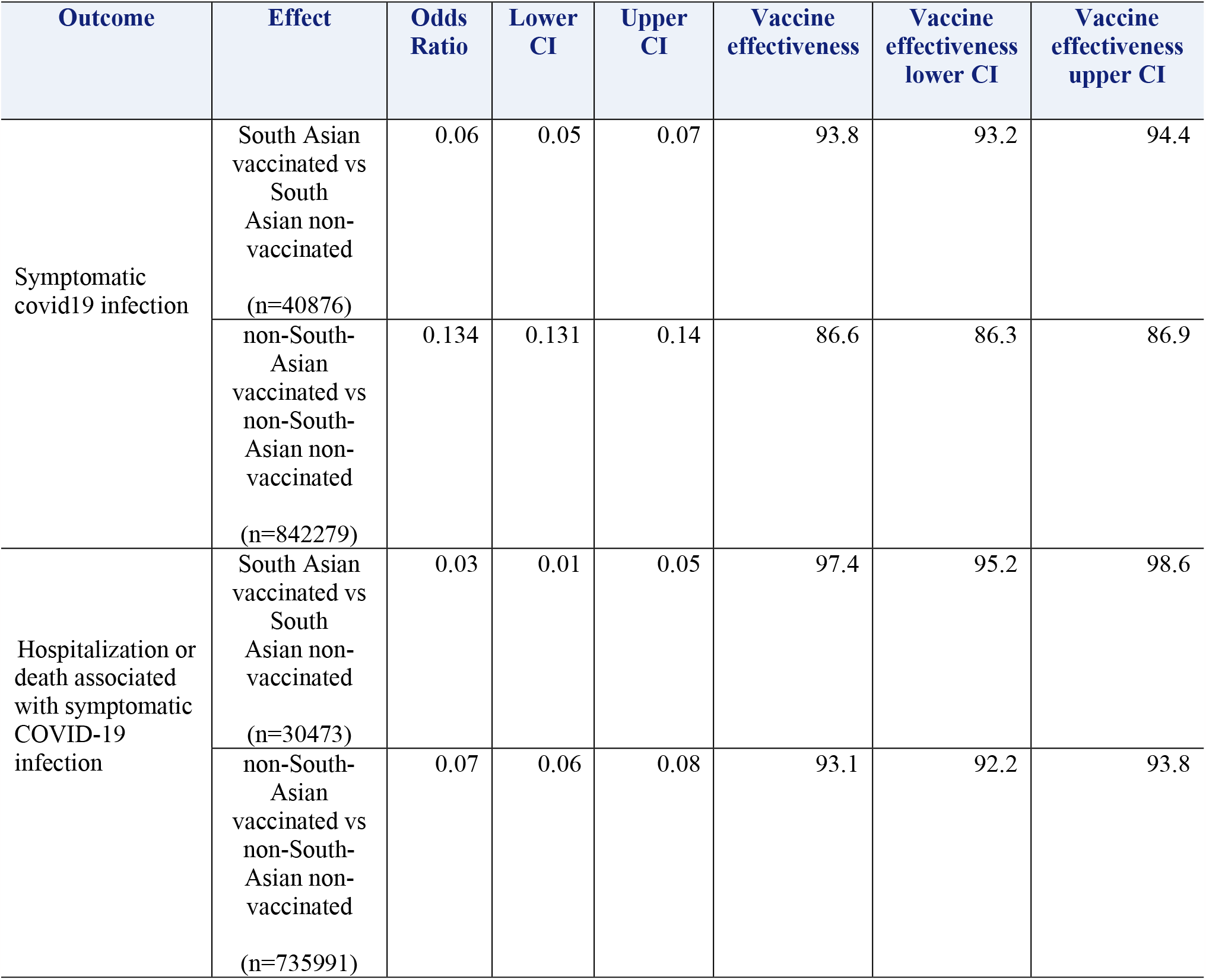
Vaccine effectiveness among South Asians and non-South Asians.

### COVID-19 infection and related hospitalizations/deaths among non-vaccinated individuals (Table 4)

**Table 4:** Adjusted logistic regression models for various outcomes in non-vaccinated South Asians, **stratified by immigration status and reason for immigration** (Referent cohort: non-South Asian non-vaccinated)

| Overall |  | Non-immigrants |  | Recent immigrant (<10 years) |  | Non-recent immigrant (>10 years) |  |
| --- | --- | --- | --- | --- | --- | --- | --- |
| Covid-19 related hospitalization or death (n=766464) | Covid-19 infection (n=883155) | Covid-19 related hospitalization or death (n=636141) | Covid-19 infection (n=712271) | Covid-19 related hospitalization or death (n=38072) | Covid-19 infection (n=50945) | Covid-19 related hospitalization or death (n=92251) | Covid-19 infection (n=119939) |
| 2.03<br>(1.9, 2.2) | 2.4<br>(2.3, 2.4) | 2.3<br>(1.9, 2.6) | 2.5<br>(2.4, 2.6) | 0.83<br>(0.6, 1.1) | 1.2<br>(1.1, 1.2) | 0.99<br>(0.9, 1.1) | 1.4<br>(1.3, 1.4) |
| Overall |  | Economic |  | Refugee |  | Family/others |  |
| Covid-19 related hospitalization or death (n=130323) | Covid-19 infection (n=170884) | Covid-19 related hospitalization or death (n=68501) | Covid-19 infection (n=84573) | Covid-19 related hospitalization or death (n=19383) | Covid-19 infection (n=28041) | Covid-19 related hospitalization or death (n= 42439) | Covid-19 infection (n=58270) |
| 0.96<br>(0.9, 1.1) | 1.30<br>(1.2, 1.3) | 0.99<br>(0.8, 1.2) | 1.2<br>(1.2, 1.3) | 0.6<br>(0.4, 0.9) | 1.1<br>(1.02, 1.3) | 1.1<br>(0.9, 1.3) | 1.5<br>(1.4, 1.6) |

Adjusted for age, sex, any comorbid condition, rural status, neighborhood income quintile

South Asian non-vaccinated individuals had a two-fold higher odds of symptomatic COVID-19 infection (aOR 2.35, 95% CI 2.3, 2.4) compared to the non-South Asian non-vaccinated cohort (referent cohort), after adjustment for multiple potential covariates. Similarly, the risk of COVID-19-related hospitalizations and deaths was 2-fold higher among non-vaccinated South Asians compared to non-vaccinated non-South Asians (aOR 2.03, 95% CI 1.9, 2.2).

### Subgroup analysis by immigration and residential status

Similar to the primary analysis, non-vaccinated South Asians compared to non-vaccinated non-South Asians had a higher odds of COVID-19 infection regardless of their immigration status or reason for immigration (Table 4).

### Subgroup analysis by reason for immigration (economic, refugee, and family/other)

Among immigrants, an increased odds of symptomatic COVID-19 infection were observed among South Asians compared to non-South Asians regardless of the reason for immigration. (Table 4). There was no difference in the risk of COVID-19 related hospitalizations and deaths among non-vaccinated South Asians and non-South Asians based on the reason of immigration, except that South Asians who immigrated to Canada as refugees had a lower risk of COVID-19 related hospitalizations and deaths compared to non-South Asian refugees (Table 4).

Similar results were obtained when further stratified analysis by reason and time since immigration (Supplemental Table 5 and Table 6).

## Discussion

In this large population-based study including close to 900,000 individuals, we show that in the third wave of the COVID pandemic in Canada, COVID-19 vaccines were effective in preventing symptomatic SARS CoV-2 infections, hospitalizations and deaths among both South Asians and non-South Asians. We also demonstrate that, among non-vaccinated individuals, South Asians had higher odds of COVID-19 infection, and an increased risk of COVID-19 hospitalizations and deaths compared to non-South Asians.

The social determinants of health are important to understand the higher impact of COVID-19 among South Asians. Some South Asian communities in Canada are vulnerable groups as they are more likely to live in multigenerational households, earn lower income compared to their education level, and are frequently employed as front-line workers, such as in transit, grocery stores, warehouses, health care, and construction (5). Due to these reasons, it is imperative to ensure adequate COVID-19 vaccine coverage among South Asians. Data from outside of Canada indicate a substantial amount of misinformation regarding the importance and effectiveness of COVID-19 vaccine in South Asian communities (10-12). Among South Asians in Canada, the top three sources of information regarding COVID-19 came from health care providers and Public Health officials, national news and traditional media sources, as well as social media (15). In addition, outside of Canada, issues such as language barriers, messaging not being tailored for the South Asian community, and lack of access to technology, were observed to be associated with lower rates of vaccine uptake and access (13). However, in Canada, South Asian advocacy groups and specialized knowledge translation strategies bridged this gap (20). Finally, ethnic communities including South Asians, were significantly underrepresented in COVID-19 vaccine trials, leading to sparse data on the effectiveness for this ethnic group (21), which may have affected vaccine confidence and rates of vaccination.

The disproportionate impact of COVID-19 among racialized groups has been confirmed by a number of studies conducted worldwide, (2, 22-26) including recent meta-analyses (25, 27). In one study which was published prior to the introduction of the COVID-19 vaccines, 18,728,893 patients were included. Black individuals (adjusted RR 2.02, 95% CI 1.67-2.44) and Asians (defined broadly, and differentiation between Chinese and South Asians was not provided) were at a higher risk of COVID-19 infections after adjusting for age, sex, and comorbid conditions (adjusted RR 1.50, 95% CI 1.24-1.83). Moreover, Asians were at higher risk of ICU admission (RR 1.97, 95% CI 1.34-2.89) compared to those of white European descent (27).

Studies evaluating the association between ethnic populations and COVID-19 in Canada are limited due to non-uniform collection of ethnicity data (28). For that reason, proxy measures of ethnicity are used. (3) In this study, we used a validated last name-based algorithm to identify South Asians living in Ontario. Our findings show that non-vaccinated South Asians have a higher risk of COVID-19 related adverse clinical outcomes. Interestingly, among South Asians, we did not find any differences in the frequency COVID-19 infection among the immigrant and non-immigrant sub-populations. These findings show that there may be several complex sociocultural factors that are in play within the South Asian community which are responsible for their increased risk of COVID-19 related adverse outcomes. It is also plausible that specific biologic pathways among South Asians account for their higher odds of COVID-19 infection (29, 30). Additionally, the potential impact of HLA and genetic factors on COVID-19 risk among different ancestral groups is also an important area for further research (31-33), and more such studies are needed among South Asians to understand their increased COVID-19 susceptibility and severity.

To our knowledge, this is the first study to evaluate the effectiveness of COVID-19 vaccines among South Asians using a test-negative design at a population level. This study design mitigates potential bias arising from differences in access to healthcare by restricting to those individuals who presented for SARS-CoV-2 testing. Despite our study’s strengths, there are some limitations that deserve mention. First, our results are generalizable only to those healthcare systems with universal healthcare coverage like Ontario. Second, there is a possibility those individuals with no information on symptoms recorded in the databases might have had symptoms at the time of testing, and those recorded as asymptomatic might have subsequently developed symptoms. Third, the last name based algorithm only allows us to identify South Asian and Chinese ethnic groups but not other ethnic communities, thus the non-South Asian group was heterogeneous by ethnicity. Fourth, we did not have data on employment, education, gender, and other social factors to further understand the increased risk of adverse outcomes among non-vaccinated South Asians. Finally, due to administrative nature of our databases, there is a potential for residual confounding and measurement error for a few variables.

## Conclusions

This study shows that COVID-19 vaccines are effective in reducing COVID-19 infections, hospitalizations and death among South Asians living in Ontario, Canada. These results should provide reassurance and foster confidence among the South Asian community regarding vaccination. Non-vaccinated South Asians have a higher odds of COVID-19 related adverse outcomes compared to non-South Asians. Future studies are needed to explain the higher risk of COVID-19 infection and worse outcomes among non-vaccinated South Asians.

## Data Availability

All data is available in the ICES databases.

## Acknowledgement

This study received funding from the COVID-19 Immune Task Force through the Public Health Agency of Canada. This study was also supported by ICES, which is funded by an annual grant from the Ontario Ministry of Health (MOH) and the Ministry of Long-Term Care (MLTC). This work was also supported by the Ontario Health Data Platform (OHDP), a Province of Ontario initiative to support Ontario’s ongoing response to COVID-19 and its related impacts. This document used data adapted from the Statistics Canada Postal Code^OM^ Conversion File, which is based on data licensed from Canada Post Corporation, and/or data adapted from the Ontario Ministry of Health Postal Code Conversion File, which contains data copied under license from ©Canada Post Corporation and Statistics Canada. Parts of this material are based on data and information compiled and provided by the MOH, the Canadian Institute for Health Information (CIHI), and Immigration Refugees and Citizenship Canada. The analyses, conclusions, opinions and statements expressed herein are solely those of the authors and do not reflect those of the funding or data sources; no endorsement is intended or should be inferred. No endorsement by the OHDP, its partners, or the Province of Ontario is intended or should be inferred.

