## Supplementary Table 1 for "COVID-19 vaccine effectiveness among South Asians in Ontario: A test-negative design population-based case-control study"

### Supplemental Table 1: List of covariates and their codes used in the study^1^

**^1^**Chung et al BMJ 2021;374:n1943 | doi: 10.1136/bmj.n1943

| Medical Condition | Definition |
| --- | --- |
| Chronic respiratory disease | Asthma:[7] An ICES-specific asthma database is used to identify patients with asthma, based on 2 or more ambulatory care visits and/or 1 or more hospitalizations. This variable is included a priori as hypothesized to be directly related to COVID-19 infection risk, as a result of its relationship to severe COVID-19 outcomes.[4]  OHIP OHIP diagnostic code: 493  DAD ICD-9 diagnostic code: 493 ICD-10 diagnostic codes: J45, J46  Chronic obstructive pulmonary disease (COPD):[8] An ICES-specific COPD database was used to identify patients with COPD, based on 1 or more ambulatory care visits and/or 1 or more hospitalizations. The algorithm to identify COPD patients was only validated in those aged 35 years or older. This variable was included a priori as hypothesized to be directly related to COVID-19 infection risk.[4]  OHIP OHIP diagnostic codes: 491, 492, 496  DAD ICD-9 diagnostic codes: 491, 492, 496 ICD-10 diagnostic codes: J41, J42, J43, J44 |
| Hypertension | An ICES-specific hypertension database is used to identify patients with hypertension, based on 1 or more DAD diagnoses or 2 or more OHIP diagnoses in a two-year period; or 1 OHIP diagnosis followed by an OHIP/DAD diagnosis within two years.[9] This variable is included *a priori* as hypothesized to be directly related to COVID-19 infection risk.[4]  DAD, SDS: ICD-9: 401, 402, 403, 404, 405; ICD-10: I10, I11, I12, I13, I15 OHIP diagnostic codes: 401, 402, 403, 404, or 405 |
| Diabetes | ODD was used to identify patients with diabetes, based on 2 OHIP diagnostic codes or 1 OHIP service code or 1 CIHI admission within 2 years.[3]  OHIP  OHIP diagnostic code: 250  OHIP service codes: Q040, K029, K030, K045, K046  CIHI-DAD, CIHI-SDS  ICD-9 diagnostic code: 250  ICD-10 diagnostic codes: E10, E11, E13, E14 |
| Immunocompromised (HIV, transplant, immunosuppressive therapy) | We included immunosuppressive conditions *a priori* as hypothesized to be directly related to COVID-19 infection risk.[4]  HIV:[11] An ICES-specific HIV database was used to identify patients with HIV, based on 3 physician claims in 3 years with OHIP diagnostic codes: 042, 043 or 044  Solid organ transplant recipients: CORRLINK is an ICES-specific database which links CORR (Canadian Organ Replacement Register) and DAD data. This database only includes patients that have received an organ transplant and does not include dialysis patients.   - For transplants before December 31, 2019: individuals are a transplant recipient if they have a treatment code of 171, where the treatment was before the index date - For transplants on/after January 1, 2020: Identify ICD-10 codes, CCI procedure codes, and OHIP feecodes from DAD, NACRS, and OHIP (codes available upon request)   Any hospitalization (any diagnosis field) with the following codes:   - Sickle-cell disease (ICD-10 D57.0 – D57.2; D57.8 OR ICD-9   282.6);   - Other immune system disorders (ICD-9 273.2, 279.0, 279.1,   279.2, 279.3, 279.8, 279.9, 289.8; ICD-10 D80, D81, D82, D83,  D84, D89; OHIP dxcode 279)   - Immunosuppressive therapy (>30 days (total days supplied) of   oral corticosteroid in the 6 months before index date; receipt of other immunocompromising drug, including antineoplastics, in the 6 months before index date)  Active cancer:  • Any of the following treatments in the past 6 months: cancer surgery (codes available upon request), radiation (if the ICD-10 code listed was Z510 in NACRS), chemotherapy (if the ICD-10 code listed was Z511 or Z512 and any evidence of cancer diagnosis in the Ontario Cancer Registry (OCR) prior to the last treatment date)  If not any of the above, individuals still classified as having cancer if they had a cancer diagnosis in OCR in the year before the index date  Allogenic/autologous bone marrow transplant recipients: We identified those who had a history of allogenic bone marrow transplant before the index date using the following combination of diagnostic codes: DAD:  CCP procedure codes = 53.0 CCI procedure codes = 1WY19, 1LZ19HHU7, 1LZ19HHU8  OHIP Feecode = Z426 |
| Chronic kidney disease (CKD) | Patients with a diagnosis of CKD in the 5 years before index date, using the following diagnostic codes [5]:  OHIP  OHIP diagnostic codes: 403, 585  CIHI NACRS, CIHI-DAD  ICD-10 diagnostic codes: E102, E112, E132, E142, I12, I13, N08, N18, N19 |
|  | Patients who were on chronic dialysis [6] in the year before index date, identified as those with at least 2 of any of the following codes in OHIP, CIHI-DAD, or CIHI-SDS separated by at least 90 days, but less than 150 days  OHIP  OHIP service codes: R849, G323, G325, G326, G860, G862, G865 G863, G866, G330, G331, G332, G333, G861, G082, G083, G085, G090, G091, G092, G093, G094, G095, G096, G294, G295, G864, H540, H740  CIHI-DAD, CIHI-SDS  CCI procedure codes: 5195, 6698  CCP procedure code: 1PZ21  CORR  Treatment codes: 060, 111, 112, 113, 121, 122, 123, 131, 132, 133, 141, 151, 152, 211, 221, 231, 241, 242, 251, 252, 311, 312, 313, 321, 322, 323, 331, 332, 333, 413, 423, 433, 443, 453 |
|  | **Exclusion criteria:**  Patients with kidney transplants [7]:  OHIP  OHIP service codes: S435, S434  CIHI-DAD  CCP procedure code: 6759  CCI procedure code: 1PC85  CORR  Treatment code: 171 plus one or more of Transplanted Organ Codes [1-3]: 10, 11, 12, 18, 19 |
| Dementia/Frailty | 1 hospitalization for dementia and/or 3 ambulatory visits for dementia, each separated by at least 30 days, within 2 years  and/or 1 prescription from ODB [9]  OHIP  OHIP diagnostic codes: 290, 331  CIHI-DAD, CIHI-SDS  ICD-9 diagnostic codes: 0461, 290.0, 290.1, 290.2, 290.3, 290.4, 294, 331.0, 331.1, 331.5  ICD-10 diagnostic codes: F00, F01, F02, F03, G30  ODB  1 prescription for a cholinesterase inhibitor  Frailty:  CIHI-DAD and OHIP databases were used to identify patients with frailty within a year before index date, based on rules that identified [10]:   1. Long-term care residence (i.e., admitted from or discharged to a nursing home after hospitalization, or location of physician billing claim was long-term care facility); 2. Receipt of palliative care; 3. Two or more domains derived from frailty scales (i.e., cognitive impairment, general health status, incontinence, falls, nutrition issues, functional performance) and health services utilization (i.e., ≥2 hospitalizations or emergency department visits, geriatrician or home care visit). |
| Chronic heart disease | Individuals are defined as having “chronic heart disease” if they have congestive heart failure (CHF), ischemic heart disease, or atrial fibrillation. The definitions for these conditions are as follows:  CHF: An ICES-derived CHF database was used to identify patients with CHF, based on 1 NACRS, DAD, SDS, or OHIP claim and a second claim (from either) in 1 year. The CHF database is limited to those aged 40 years or older. This variable was included a priori as hypothesized to be directly related to COVID-19 infection risk.[4]  OHIP: 428 DAD, SDS: ICD-9: 428, ICD-10: I500, I501, I509  Cardiac ischemic disease:[5] Any comorbidity in the past 5 years (DAD, any diagnosis field) or history of procedure in past 20 years (DAD, SDS), of the following:  Comorbidity (DAD, any diagnosis in the past 5 years): Angina: ICD-10: I20  Chronic Ischemic Heart Disease: ICD-10: I25; Myocardial infarction: ICD-10: I21, I22  Procedure (DAD & SDS): Coronary Artery Bypass Grafting: CCI procedure codes: 1IJ76 CCP procedure codes: 481  Percutaneous Coronary Intervention: CCI procedure codes: 1IJ50, 1IJ54, 1IJ57GQ CCP procedure codes: 4802, 4803  Atrial fibrillation:[6] Individuals with 1 hospitalization or 4 MD visits within a year in the past 5 years with the following codes: ICD-9: 427.31, 427.32 ICD-10: I48 OHIP dxcode: 427 |
| History of TIA or Acute Ischemic Stroke | Transient Ischemic Attack:  CIHI-DAD and CIHI-NACRS were used to identify patients with a history of a transient ischemic attack, based on at least 1 hospitalization or ED visit with a diagnosis coded with one of the following codes:  ICD-9 diagnostic codes: 435, 3623  ICD-10 diagnostic codes: G450, G451, G452, G453, G458, G459, H340  Acute Ischemic Stroke [12]:  CIHI-DAD was used to identify patients with a history of acute ischemic stroke, based on at least 1 hospitalization with a main diagnosis coded with one of the following codes:  ICD-9 diagnostic codes: 434, 436  ICD-10 diagnostic codes: I63 (excluding I63.6), I64, H34.1 |
| Autoimmune disease | Individuals considered to have autoimmune disease if they had any of the following:   - Rheumatoid arthritis (identified in the Ontario Rheumatoid Arthritis Database)[13,14] - Inflammatory bowel disease (identified in the Ontario Crohn’s and Colitis Cohort)[15,16] - Psoriasis/psoriatic arthritis[17]   o Psoriasis: 1 hospitalization or 3 physician billings prior to  index date. DAD: ICD-9: 696.1, 696.8; ICD-10: L40.0, L40.1, L40.2, L40.3, L40.4, L40.8, L40.9. OHIP: dxcode = 696  o Psoriatic arthritis: 1 hospitalization or: (3 physician billings for psoriatic arthritis + 1 billing for psoriasis [696]). DAD: ICD-9: 696.0; ICD-10: L40.5, M07.0, M07.1, M07.2, M07.3, M09.0. OHIP: dxcode = 721 (at least one of these billings must be billed by a rheumatologist).  • Multiple sclerosis[18] o Individuals with one hospitalization or 5 physician billings over 2 years. DAD: ICD-9: 340; ICD-10: G35. OHIP: dxcode = 340 |
| Advanced liver disease | We included advanced liver disease *a priori* as hypothesized to be directly related to COVID-19 infection risk.[4]  Defined using the Cirrhosis Algorithm 9 [from [21]]: Two or more physician visits (diagnosis code 571), or one or more hospital diagnosis of cirrhosis, using the following diagnostic codes: ICD-9 : 456.1, 571.2, 571.5  ICD-10: I85.9, I98.2, K70.3,K71.7, K74.6  Defined using the Decompensated Cirrhosis Algorithm 5 (from [21]): One or more physician visits with diagnosis code 571 and (one or more hospital diagnosis or one or more procedure), using the following diagnostic codes:  ICD-9: 456.0, 456.2, 572.2, 572.3, 572.4, 782.4, 789.5l; ICD-10: I85.0, I86.4, I98.20, I98.3, K721, K729, K76.6, K76.7, R17, R18  CCI: 1.NA.13.BA-FA, 1.NA.13.BA-X7, 1.NA.13.BA-BD, 1.KQ.76GP-NR, 1.OT.52.HA  CCP: 1006, 6691 OHIP: J057, Z591 |
