## Supplementary Table 2 for "COVID-19 vaccine effectiveness among South Asians in Ontario: A test-negative design population-based case-control study"

### Supplemental Table 2: Cohort Creation Process

| **Step** | **Description** | **# of individuals** |
| --- | --- | --- |
| case group |  |  |
| 1 | get all patients who had symptoms at the covid19 test (from olisc19.covid19_symptoms) | 1,979,889 |
| 2 | get all patients with positive covid19 test (from c19intgr.c19intgr) | 1,307,098 |
| 3 | get patients with positive covid19 tests and symptoms | 507,269 |
| 4 | get patients with positive covid-19 tests from 2020-12-14 to 2021-11-15 and symptoms | 153,927 |
| control group |  |  |
| 5 | get all patients who ever had negative/indeterminate covid-19 test | 7,178,821 |
| 6 | get patients with negative covid19 test and symptoms | 1,840,670 |
| 7 | remove patients who had negative test but also ever had positive test | 1,464,959 |
| 8 | get patients with negative covid-19 tests from 2020-12-14 to 2021-11-15 and symptoms | 1,096,777 |
| combine |  |  |
| 9 | combine case and control groups | 1,250,704 |
| 10 | age must be 18+, with valid gender, reside in Ontario at index date, delete death before index date | 906,293 |
| 11 | remove patients living in long term care (LTC) | 886,325 |
| 12 | exclude patients with positive covid-19 test before 2020-12-13 | 885,526 |
| 13 | remove patients who was not covered by OHIP at index date | 883,155 |
| **final cohort** |  | **883,155** |
