## Supplemental Tables 3 and 4 for "COVID-19 vaccine effectiveness among South Asians in Ontario: A test-negative design population-based case-control study"

### Supplemental Table 4: Baseline comorbidities of the overall cohort stratified by covid-19 test results

| **Comorbidities** | **People with negative covid-19 test and symptoms** | **People with positive covid-19 test and symptoms** | **STANDARDIZED DIFFERENCE** |
| --- | --- | --- | --- |
| Any comorbidity | 360,271 (47.6%) | 54,259 (43.1%) | 0.09 |
| Chronic respiratory disease (Asthma, COPD) | 196,796 (26.0%) | 25,742 (20.4%) | 0.13 |
| Asthma | 157,282 (20.8%) | 21,724 (17.2%) | 0.09 |
| COPD | 61,356 (8.1%) | 6,286 (5.0%) | 0.13 |
| Chronic heart disease (CHF, IHD, AF) | 56,264 (7.4%) | 6,882 (5.5%) | 0.08 |
| Heart failure | 21,851 (2.9%) | 2,455 (1.9%) | 0.06 |
| Ischemic heart disease | 22,332 (2.9%) | 2,727 (2.2%) | 0.05 |
| Atrial fibrillation | 32,244 (4.3%) | 3,875 (3.1%) | 0.06 |
| Angina | 2,564 (0.3%) | 270 (0.2%) | 0.02 |
| Chronic ischemic heart disease | 11,812 (1.6%) | 1,426 (1.1%) | 0.04 |
| MI | 6,727 (0.9%) | 775 (0.6%) | 0.03 |
| CABG | 5,845 (0.8%) | 733 (0.6%) | 0.02 |
| PCI | 13,097 (1.7%) | 1,700 (1.3%) | 0.03 |
| Hypertension | 162,899 (21.5%) | 25,793 (20.5%) | 0.03 |
| Diabetes | 82,839 (10.9%) | 16,053 (12.7%) | 0.06 |
| Immunocompromised disorders | 39,925 (5.3%) | 4,180 (3.3%) | 0.1 |
| HIV | 1,355 (0.2%) | 252 (0.2%) | 0 |
| Solid organ transplant or bone marrow transplant | 2,099 (0.3%) | 286 (0.2%) | 0.01 |
| Sickle cell anemia | 224 (0.0%) | 60 (0.0%) | 0.01 |
| Other immune system disorders | 13,525 (1.8%) | 1,584 (1.3%) | 0.04 |
| immunosuppressive therapy | 14,113 (1.9%) | 1,493 (1.2%) | 0.06 |
| Treatment in past 6m or recent diagnosis in OCR | 14,095 (1.9%) | 1,054 (0.8%) | 0.09 |
| Transplant in CORR or DAD/NACRS/OHIP | 1,237 (0.2%) | 208 (0.2%) | 0 |
| bone marrow/stem cell transplant | 871 (0.1%) | 78 (0.1%) | 0.02 |
| Autoimmune disease | 32,862 (4.3%) | 4,173 (3.3%) | 0.05 |
| Rheumatoid arthritis | 9,999 (1.3%) | 1,267 (1.0%) | 0.03 |
| Inflammatory bowel disease | 5,371 (0.7%) | 623 (0.5%) | 0.03 |
| Psoriasis | 15,168 (2.0%) | 2,033 (1.6%) | 0.03 |
| Psoriatic arthritis | 1,793 (0.2%) | 213 (0.2%) | 0.02 |
| Multiple sclerosis | 3,391 (0.4%) | 369 (0.3%) | 0.03 |
| Chronic kidney disease or dialysis | 23,887 (3.2%) | 3,367 (2.7%) | 0.03 |
| Chronic kidney disease | 23,886 (3.2%) | 3,366 (2.7%) | 0.03 |
| Dialysis in each of 3 months prior to indexdate | 1,568 (0.2%) | 216 (0.2%) | 0.01 |
| Advanced liver disease | 8,065 (1.1%) | 1,019 (0.8%) | 0.03 |
| Cirrhosis | 7,597 (1.0%) | 962 (0.8%) | 0.03 |
| Decompensated cirrhosis | 1,291 (0.2%) | 152 (0.1%) | 0.01 |
| Dementia | 11,506 (1.5%) | 1,417 (1.1%) | 0.03 |
| Transient ischemic attack or stroke | 11,079 (1.5%) | 1,229 (1.0%) | 0.04 |
| Transient ischemic attack (TIA) | 6,896 (0.9%) | 680 (0.5%) | 0.04 |
| Acute ischemic stroke | 5,195 (0.7%) | 641 (0.5%) | 0.02 |
