## Supplemental Tables 5 and 6 for "COVID-19 vaccine effectiveness among South Asians in Ontario: A test-negative design population-based case-control study"

### **Supplemental Table 5:** Adjusted logistic regression models for symptomatic covid-19 infection in non-vaccinated South Asians, stratified by reason of immigration and years (Referent cohort: non-South Asian non-vaccinated)

| **cohort** | **status** | **Odds Ratio** | **Lower CI** | **Upper CI** | **P value** | **number of patients** |
| --- | --- | --- | --- | --- | --- | --- |
| overall | Recent  immigrant (<10  years) | 1.2 | 1.1 | 1.2 | <.0001 | 50945 |
| overall | Non-recent  immigrant  (>=10 years) | 1.4 | 1.3 | 1.4 | <.0001 | 119939 |
| Economic | Recent  immigrant (<10  years) | 1.2 | 1.1 | 1.3 | <.0001 | 26315 |
| Economic | Non-recent  immigrant  (>=10 years) | 1.3 | 1.2 | 1.4 | <.0001 | 58258 |
| Refugee | Recent  immigrant (<10  years) | 1.3 | 0.9 | 1.4 | 0.1 | 7933 |
| Refugee | Non-recent  immigrant  (>=10 years) | 1.2 | 1.05 | 1.3 | 0.005 | 20108 |
| Family/other | Recent  immigrant (<10  years) | 1.6 | 1.5 | 1.8 | <.0001 | 16697 |
| Family/other | Non-recent  immigrant  (>=10 years) | 1.5 | 1.4 | 1.6 | <.0001 | 41573 |

Adjusted for age, sex, any comorbid condition, rural status, neighborhood income quintile
