## Supplemental Tables 5 and 6 for "COVID-19 vaccine effectiveness among South Asians in Ontario: A test-negative design population-based case-control study"

### **Supplemental Table 6:** Adjusted logistic regression models for covid-19 related hospitalization or death, stratified by type of immigration and years in non-vaccinated South Asians (Referent cohort: non-South Asian non-Vaccinated)

| **Cohort** | **status** | **Odds Ratio** | **Lower CI** | **Upper CI** | **P value** | **number of patients** |
| --- | --- | --- | --- | --- | --- | --- |
| overall | Recent immigrant (<10 years) | 0.8 | 0.6 | 1.1 | 0.2 | 38072 |
| overall | Non-recent immigrant (>=10 years) | 0.99 | 0.9 | 1.1 | 0.9 | 92251 |
| Economic | Recent immigrant (<10 years) | 0.9 | 0.6 | 1.4 | 0.6 | 20860 |
| Economic | Non-recent immigrant (>=10 years) | 1.1 | 0.9 | 1.3 | 0.6 | 47641 |
| Refugee | Recent immigrant (<10 years) | 0.4 | 0.1 | 1.8 | 0.2 | 5010 |
| Refugee | Non-recent immigrant (>=10 years) | 0.7 | 0.4 | 0.9 | 0.03 | 14373 |
| Family/other | Recent immigrant (<10 years) | 1.3 | 0.9 | 1.9 | 0.2 | 12202 |
| Family/other | Non-recent immigrant (>=10 years) | 1.1 | 0.9 | 1.3 | 0.5 | 30237 |

Adjusted for age, sex, any comorbid condition, rural status, neighborhood income quintile
