## Supplementary figures and images for "COVID-19 vaccine effectiveness among South Asians in Ontario: A test-negative design population-based case-control study"

### Supplemental Figure 1

#### Supplemental Figure 1: Participant flow diagram

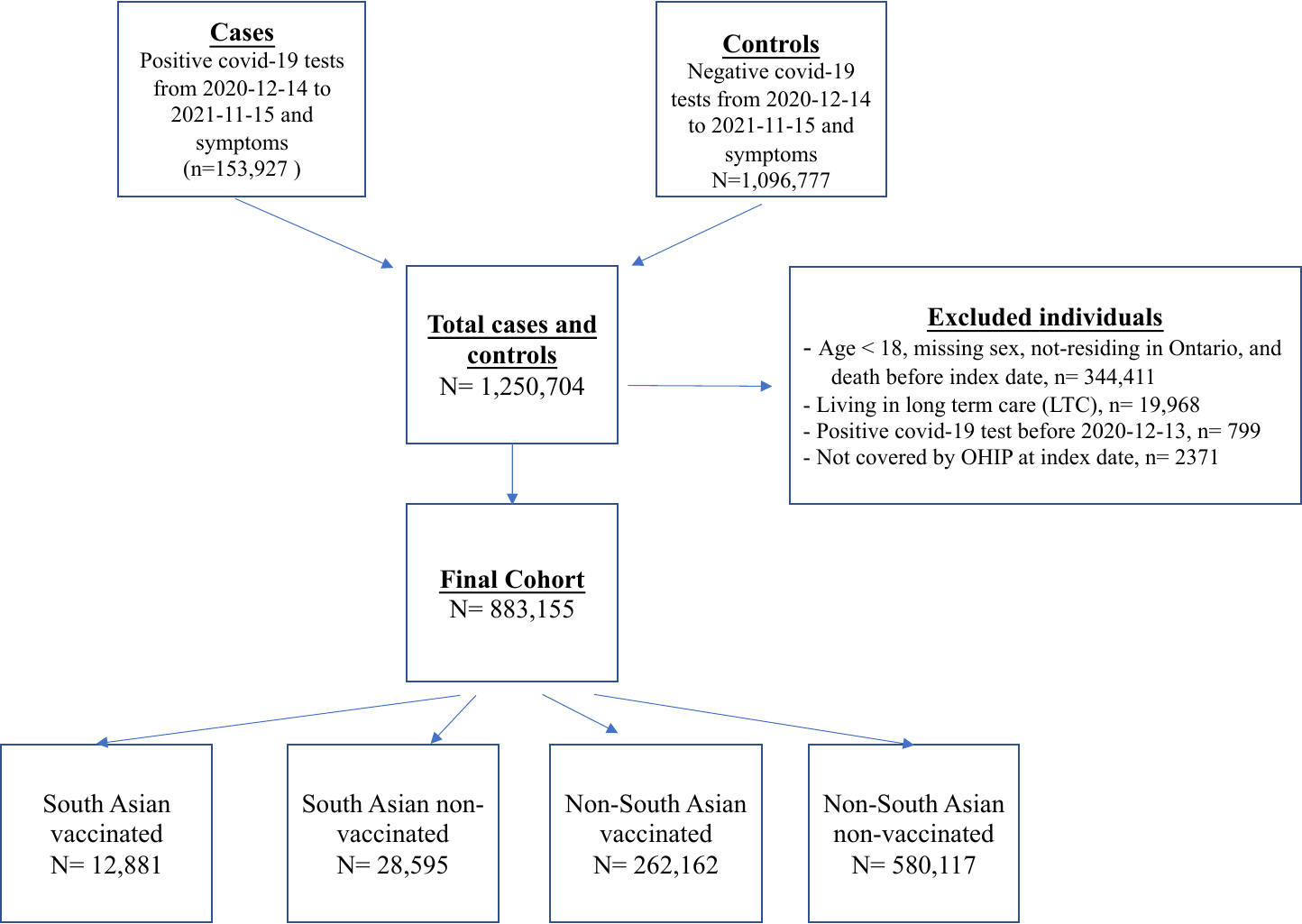
